# Patient Centric Summarization of Radiology Findings using Large Language Models

**DOI:** 10.1101/2024.02.01.24302145

**Authors:** Amara Tariq, Sam Fathizadeh, Gokul Ramaswamy, Shubham Trivedi, Aisha Urooj, Nelly Tan, Matthew T. Stib, Bhavik N. Patel, Imon Banerjee

## Abstract

**Objective:** Develop automated AI models for patient-sensitive summarization of radiology reports. Level of medical education or socio-economic background of a patient may dictate their level of understanding of medical jargon. Inability to understand primary findings from a radiology report may lead to unnecessary anxiety among patients or result in missed follow up.

**Materials and Methods:** Computed tomography exams of chest were selected as a use-case for this study. Approximately 7K chest CT reports were collected from Mayo Clinic Enterprise. Summarization model was built on the T5 large language model (LLM) as its text-to-text transfer architecture is intuitively suited for abstractive text summarization, resulting in a model size of ~0.77B. Noisy groundtruth for model training was collected by prompting LLaMA 13B model.

**Results:** We recruited both experts (board-certified radiologists) and laymen to manually evaluate summaries generated by model. Model-generated summaries rarely missed information as marked by majority opinion of radiologists. Laymen indicated 63% improvement in their understanding by reading layman summaries generated by the model. Comparative study with zero-shot performance of LLaMA indicated that LLaMA hallucinated and missed information 3 and 4 times more often, respectively, than the proposed model.

**Discussion:** The proposed patient-sensitive summarization model can generate summaries for radiology reports understandable by patients with vastly different levels of medical knowledge. In addition, task-specific training allows for more reliable performance compared to much larger off-the-shelf models.

**Conclusions:** The proposed model could improve adherence to follow up treatment suggested by radiology reports by increasing patients’ level of understanding of these reports.

## INTRODUCTION

The 21st Century Cures Act’s mandate for immediate release of electronic health information (EHR) facilitated patients’ access to their radiology reports, often before referring physicians can explain to patients their radiological findings’ significance. These diagnostic and procedural reports are composed of complex jargon and non-grammatical fragments, with less than 4% of reports at the 8th grade reading level of the average United States adult [1]. In today’s practice, medical records in the EHR such as radiology reports are largely constructed for provider-to-provider communication and for ICD/CPT coding for billing. These medical reports are now being increasingly accessed by patients. This can contribute to significant patient confusion and anxiety, which in turn can lead to confusion about one’s medical care and may contribute to lack of adherence to follow-up or treatment[2], [3]. Moreover, complex language within the report may contribute to lack of follow up for incidental and other findings suggested within the report. It has been shown that only 50% of recommended follow up is performed[4]. While some work has been done to study the effect of patient-level factors on adherence to follow up recommendation [5], negligible research effort has been directed towards making radiology reports readable to patients by taking patient-level factors into account.

Several studies in the past have established a direct link between patients’ understanding of their medical information with adherence to recommended prevention and treatment processes, better clinical outcomes, better patient safety within hospitals, and less health care utilization[10]. Radiology reports are an integral part of medical information but are traditionally hard to understand by the patients, even the impression, as they are mainly written for communication between radiologists and clinical specialists. Large amount of Natural Language Processing (NLP) research effort has been directed towards automated generation of radiology reports, often with radiology images as input[11], [12], automated writing of the Impression section[13]–[15] with focus on factual correctness [16]. While previously recurrent neural networks were specifically trained for this task, LLMs are now being tested on this application as zero-shot learners[17], [18]. However, most of these models are not designed with patients in mind, and hence, their output is not sensitive to any patient characteristics. We attempted to solve this issue by providing the proposed model information about the patient so that the model’s output was tailored to the patient. This additional step will ensure that communication between radiologists and clinical specialists remains unaffected as the original radiology report writing standards are not altered for increasing patients’ understanding level by potentially compromising information delivery to clinical specialists. Building an automated model to do this job will ensure that radiologists’ workload does not increase in any manner.

We hypothesized that an ‘understandable’ radiology report summary will look different for someone with a high-school diploma compared to a clinical professional. Hence, any radiology report simplification model must be conditioned on the patient. While general-purpose large languages like GPT and LLaMA may be used as zero-shot learners for this task [6], studies have shown that their off-the-shelf use in sensitive fields like medicine is inadvisable given inconsistency in their performance and their propensity to “hallucinate” information[7]–[9]. To reliably achieve the goal of enhancing patient understanding of radiology reports in a safe and sensitive manner, we developed and trained an innovative model built based on a publicly available and relatively “smaller” LLM whereas the training data for this novel task was curated using the largest publicly available LLM in addition to manual filtering effort. This approach of using larger LLM in the inference model and fine tuning a smaller LLM allowed us to curtail computational costs while achieving high task-based performance.

Large language models (LLM) are huge models with 100s of billions of parameters trained on extremely large textual datasets like text crawled from the world wide web, often in self-supervised manner with tasks like masked token prediction or next sentence prediction. LLMs have shown astounding capabilities in solving many NLP tasks. Their biggest advantage is their performance as zero-shot learners promising the availability of a universal language model - one model that can perform any language based extraction, retrieval or even reasoning based tasks[19]–[23]. However, this approach may not be suitable for application in every domain, Sensitive domains like medicine require precise use of language in a consistent manner. LLMs have displayed trends of inconsistency in performance - different output for the same input - and hallucination where the model seems to “imagine” information that does not exist [7]–[9]. Additional problems arise when the model is used to make recommendations and those recommendations are not concordant with current clinical standards as set by clinical expert bodies like ASCO and NCCN[24]. In the case of communicating radiologic findings with patients, these trends may result in inconsistent, inaccurate, or misleading information to be disseminated.

While extremely large language models have been able to show astounding generalization capacity, training of such models requires extremely large training datasets and huge amounts of computational resources. We argue that there is a balance to strike between computational resources and performance, especially when the model is targeted for a specific use-case and not intended to be used for unrelated applications. Our use case falls within this scenario. Therefore, we experimented with a relatively smaller LLM and showed that it was able to achieve reliable performance for the given task while curtailing the need for computational resources. Our novel use case had no available verified training data - layman summary of the radiology findings. Thus, we decided to make use of the ability of much larger LLMs to generalize to novel tasks by generating noisy ground truth for training our model. We later used manual filtering techniques to dropout low-quality data points. This approach limited the amount of manual effort needed as much larger time and effort would have been needed to generate groundtruth data from scratch. In addition, larger LLMs were only used in inference mode without finetuning or training thus limiting the number of computational resources used. Two-pronged user and expert studies were conducted to evaluate the performance of our model. The results establish the efficacy of our model in terms of generating understandable and accurate summaries sensitive to the patients’ levels of medical knowledge.

## MATERIALS AND METHODS

The proposed problem requires generation of two versions summary for each radiology report - 1) technical summary meant for patients or referring physicians with a high level of understanding of medical jargon, 2) layman summary meant for people with limited knowledge of medical terminology. In Fig 1, we presented the overall framework for generating two versions of summary of findings documented within radiology reports which contains the three primary modules - (i) section segmentation; (ii) noisy data generation for layman summary; (iii) two-step large language model fine-tuning; and (iv) user evaluation to evaluate the quality of both technical and layman summaries.

**Figure 1.**
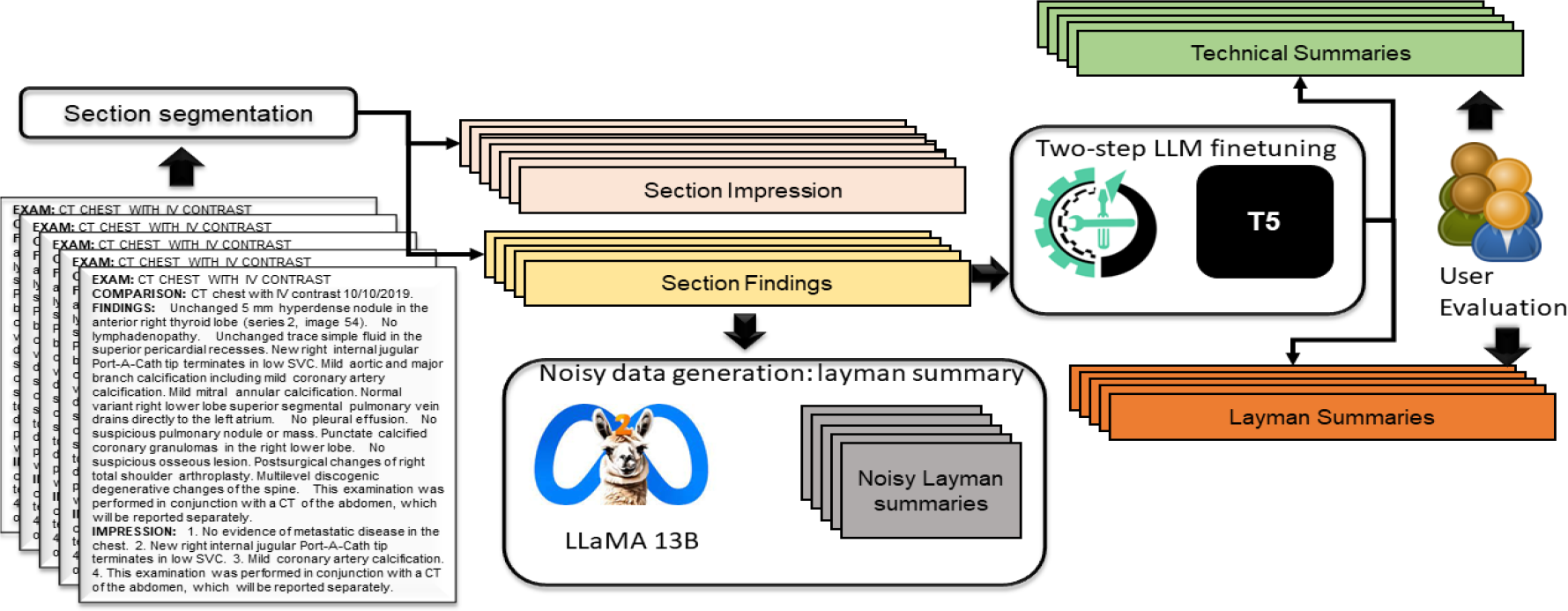
Overall pipeline of generating two different versions of summary of radiology reports - technical summary (similar to “impression” section) and layman summary.

### Section segmentation

We utilize previously developed NLP methods to parse the clinical history, imaging protocol, findings, and impression sections of the radiology reports using section segmentation based on the header[25]. To generalize the section segmentation across multiple institutions, we extracted all the variations of the headers using a similar word list generated by Word2Vec language model trained on 3M radiology reports from Emory University Hospital. Such a non-contextual language model generates a similar word list only by reflecting co-occurrence statistics which is sufficient for capturing header variations between reports given that such words appear in similar context. We computed the similar word list for each header by intersecting the list of other headers. For example, similar wordlists for the ‘clinical history:’ section include: ‘indications:’, ‘history:’, ‘patient history:’, ‘reason for exam’, ‘reason for order’. In the later sections, we only utilize the ‘finding’ and ‘impression’ section from the original report.

### Noisy data generation

The most challenging aspect of radiology finding simplification is the lack of robust groundtruth, i.e., patient understandable summary, for model training while we plan to use the original ‘impression’ section from the radiology reports. To the best of our knowledge, no dataset of radiology reports and their corresponding patient-understandable summaries exists, and generating those summaries manually is extremely expensive and variable based on manual experience. We decided to meet this challenge through the use of weak ground truth generated by publicly available large language models (LLM) - LLaMA 13B model [26] where we instruct the model to generate patient-understandable summaries given the radiologic findings as input (see Fig 2 for the exact prompt). In our study, this summary generation process served another alternative purpose - for the given radiology report summarization task, study of zero-shot performance of very large state-of-the-art LLMs - LLaMA 13B. However, we call these generative summaries - ‘weak groundtruth’ since tendencies of foundational model *zero shot* summaries are known to have inconsistency and hallucination which puts limits on their off-the-shelf use for sensitive applications like communication with patients. Hallucinated information may increase patient anxiety. Inconsistency in model response may increase confusion among patients.

**Figure 2.**
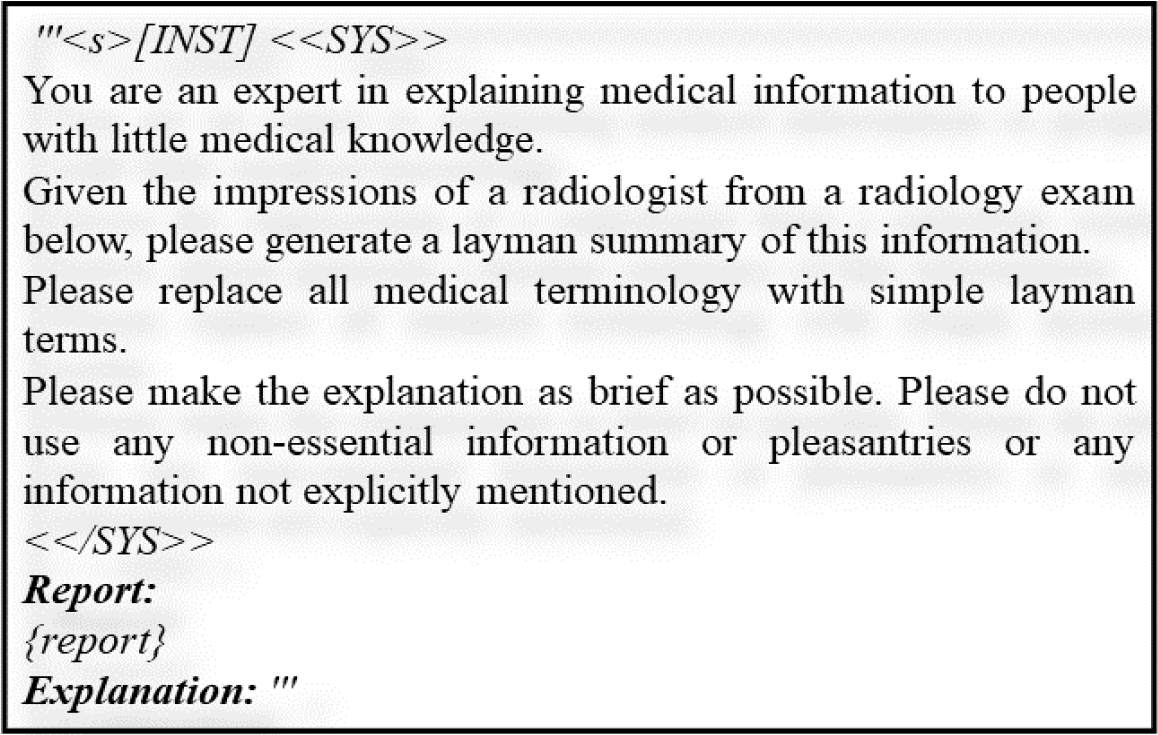
Sample prompt for generating noisy ground truth for layman summary.

### Two-step LLM fine-tuning

For the specialized task of radiology simplification, we opted to fully fine-tuned LLM models which require availability of labeled training data. However, we designed a two-step training process - **(step 1)**: technical summary generation with <input: finding and output: original impression section> as labeled pair; **(step 2):** user sensitive radiology report summarization with <input: finding and output: LLaMA generated layman summary> as labeled pair with weak ground truth; Our LLM summarization model was based on the architecture and initial weights of the text-to-text transfer transformer (T5) model publicly released by Google [27]. We used their “large” version consisting of 770 million parameters which is considerably smaller than off-the-shelf open-source popular LLMs like GPT-2 (1.5B) and LLaMA (7B, 13B, and 65B parameter version), and thus it should be comparatively easy to fine-tune with limited data. The selection was also based on intuitive adaptability of text-to-text transfer architecture for “transferring” report to summaries as displayed by the models original target tasks which included summarization [27].

Given the fact that the desired model is supposed to be sensitive to the user’s level of medical knowledge, we designed the modeling framework to utilize instruction-based prompts for providing information about a user’s level of medical knowledge in the form of free-text encoding which does not require any change in the model architecture. For practical purposes, the current version of the proposed model expects two prompts for generating two different summaries of the same report, i.e., layman and expert levels of knowledge.

### User-centric evaluation (radiologist and users with non-medical background)

As discussed earlier, the groundtruth labeled pair does not exist for the layman summary. Thus, we decided to formulate two-pronged strategy for true evaluation of the summaries generated by the model to evaluate - i) model generated summary especially layman level summary is understandable by the patients with little-to-moderate level of medical knowledge, and ii) model generated both layman and technical summaries are providing correct and complete information without hallucination.

*In evaluation I*, 3 users with limited medical knowledge and different education levels were asked to grade on Likert scale [28] within range of 1 to 5 with 5 being the perfect score if the layman summary generated by the model is understandable or not. *In evaluation II*, the 3 expert board certified radiologists were asked to read both the layman and expert level summaries and fill out a detailed survey based on three aspects; a) *missing information:* is there any information missing that should have been communicated to the patient (evaluated on binary scale: Yes/No), b) *hallucination:* is there any “hallucinated’’ information in the model generated summary (evaluated on binary scale: Yes/No), and c) *linguistic accuracy:* semantic quality of the summary language (evaluated on Likert scale within range of 1 to 5 with 5 being the perfect score). The results of both studies are reported in Table 4.

## Results

### Dataset

With the approval of Mayo Clinic Institutional Review Board (IRB), we chose a commonly performed diagnostic exam of chest CT as the primary use-case for experiments. Approximately 120K chest CTs are performed each year in Mayo Clinic for new diagnosis and/or tracking progression of a variety of issues including chest infection. heart and lung problems, blocked arteries, congestive heart failure, lung cancer, pulmonary embolism, damage to lymph nodes, muscle and bone disorders, bone tumors and fractures, and internal bleeding. While inability to identify incidental findings may lead to delayed followup, misunderstandings regarding critical issues like malignant lesions in lungs may lead to increased anxiety among patients. We collected 6970 chest CT reports from Mayo Clinic for exams performed between 2013 and 2022. The following table described major characteristics for these reports, including distribution of findings.

### Quantitative Evaluation

To evaluate the performance quantitatively, we first ran experiments with progressively larger filtered training sets; 2k, 3k, 4k, and 6k after manually filtering out low quality ground truth summaries generated by LLaMA (e.g., incomplete information, hallucinations, linguistic error). Model overfitted severely for dataset sizes up to 3K. Larger training set (>3K) sizes resulted in better learning curves of the training process as depicted in Figure 3 (training curves from all 4 experiments).

**Figure 3:**
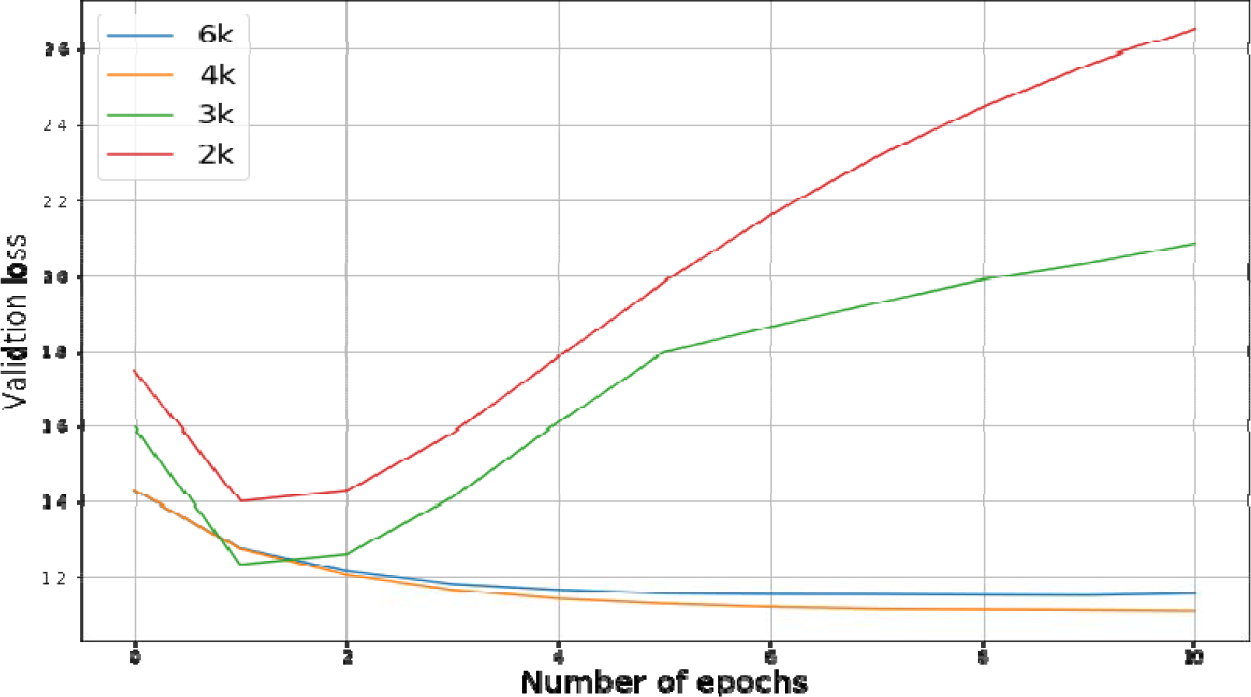
Effects of training dataset size on cross entropy-based validation losses during training of the proposed summarization model. Training datasets sizes (shown in legend) were progressively increased as this process required manual effort.

Once the model training was stabilized, we manually compared the performance of the proposed model with the use of LLaMA as zero-shot learner for layman level summary generation over randomly sampled subset of 100 reports in terms of hallucination and missing information. Table 2 presented the comparative performance where T5 model (less no. of trainable parameters) with task specific fine-tuning resulted in less hallucination and missing information and Table 3 shows some actual examples generated by LLAMA and our trained model.

**Table 1:** Study cohort characteristics. Findings are not mutually exclusive.

|  |  |
| --- | --- |
| **Total number of reports** | 6970 |
| **Years of study** | 2013 - 2022 |
| **Mean length of finding section** | 150 +/- 65 words |
| **Mean length of impression** | 33 +/- 26 words |
| **Patient Characteristics** | Age: 66.7+/-10.2 year<br>Gender: 54% male, 46% female<br>Race: 91.6% Caucasian, 3.5% African American, 1.4% Asian |
| **Major findings** | Lung nodules: 5276<br>Cardiovascular issues: 4617<br>Fractures: 771<br>Metastatic cancer: 432<br>Stable exam: 272 |

**Table 2:**
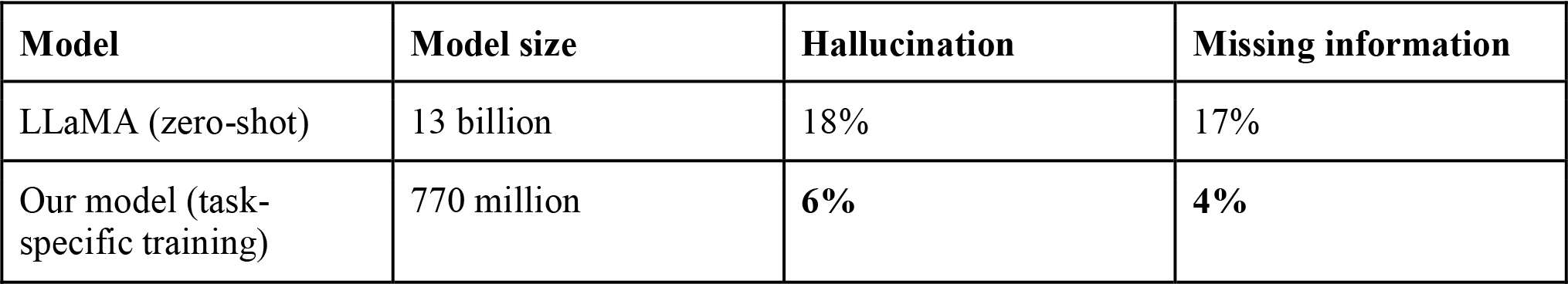
Comparison between the performance of the proposed model and zero-shot performance of LLaMA for layman summary generation. Optimal performance highlighted in bold.

| Model | Model size | Hallucination | Missing information |
| --- | --- | --- | --- |
| LLaMA (zero-shot) | 13 billion | 18% | 17% |
| Our model (task-specific training) | 770 million | **6%** | **4%** |

**Table 3:** Sample outputs of LLaMA and the domain-specific fine-tuned proposed model. ‘…’ represents additional text which is not added to preserve readability.

| Original Finding Section | LLaMA-generated layman summary | Layman summary- Our model | comment |
| --- | --- | --- | --- |
| Two new sub-3 mm micronodules in the right middle lobe ... groundglass suggesting an infectious/inflammatory etiology. A 3 mm nodule in the central left upper lobe .... Coronary artery calcification. Trace pericardial fluid. ... | 1. The radiologist found two small spots in the right middle lobe of the lung that may be caused by an infection or inflammation, but it's not clear and needs to be monitored. 2. The rest of the <u>chest X-ray</u> looks the same as it did in [DATE] | The radiologist found two small, new spots on the right side of your lung that are not yet clear what they are. They may be caused by an infection or inflammation. The radiologist also found some cloudy areas in your lungs that could be caused by an infection or inflammation. | LLaMA <u>hallucinated the report originated from</u> “X-ray” exam |
| a few scattered tiny solid pulmonary nodules in the | The radiologist found a small increase in the | The radiologist found a small area of thickening | LLaMA <u>missed</u> lung nodules, |
| upper lobes are unchanged since [DATE] and should be benign ... mild upper lung predominant centrilobular emphysema. mild biapical scarring. mild dependent atelectasis. new slight peripheral interlobular septal thickening in the basal lower lobes posteriorly .... | thickness of the tissue in the lower lobes of the lungs, specifically in the back of the lungs. This could be caused by a reaction to a medication or slight fluid buildup in the lungs. | in one of the lower lobes of the lungs, which could be caused by an infection or inflammation. The radiologist also found some small, solid nodules in the upper lobes of the lungs, which are stable and should be benign. | Our model included a description of these nodules in the summary. |
| no appreciable change in a slightly irregular 4 mm lateral left upper lobe nodule on .... sub-solid nodules measuring 5 mm or less, .... stable clustered micronodularity in the left lower lobe peripherally and right upper lobe medially . stable tiny nodule/lymph node near the right minor fissure . calcified granulomas. tiny foci of endobronchial mucus plugging scattered in both lungs. ... | This examination was performed to evaluate the lymph nodes in the neck and the soft tissues of the neck.<br>Impression:<br>1. there is a 2.0 cm short axis mass in the right level II lymph node. 2. there is a 1.5 cm short axis mass in the left level II lymph node. 3. there is a 1.0 cm short axis mass in the right level III lymph node. 4. there is a 0.5 cm short axis mass in the left level III lymph node." | The radiologist found no signs of cancer spreading to the lungs. | LLaMA <u>hallucinated</u> masses and their sizes.<br>Our model <u>missed</u> that there was no change in lung nodules. |
| central airways are patent. fat-containing right posterior diaphragmatic bochdalek hernia. small hiatal hernia. heart and pericardium: left atrial enlargement; .... changes of the spine. please see mri abdomen report from today for further description of abdominal findings including known pancreatic mass with biliary stent in place and pneumobilia. | Here is the layman summary of the report:<br>The MRI scan of your abdomen shows:<br>* Your liver is slightly larger than normal and there are no signs of cancer in it. * Your spleen is also slightly larger than normal and there are no signs of cancer in it. * Your pancreas is ... | The radiologist found no signs of cancer spreading to the lungs. | LLaMA <u>hallucinated</u> MRI scan report and abdominal findings. |
| ... focal sclerosis of the anterolateral left sixth rib is unchanged .... mildly enlarged 11 mm | The radiologist found some small spots on the lungs that are not cancerous. Some of these | The radiologist found a new area of hardening on the left side of the sixth rib, <u>which could be a sign</u> | Our model <u>hallucinated</u> possible cause of focal |
| paraesophageal lymph node is unchanged since ... increased small partially loculated left greater than right pleural effusions. there is mid and upper lung predominant interstitial abnormality characterized by subpleural reticulation. .... many of the pre-existing bilateral pulmonary nodules measuring up to 6 mm are unchanged since ... | spots are the same size as before, while others have grown a bit. The doctor will use these images as a baseline to compare future scans. There is also some fluid buildup around the lungs and an enlarged lymph node near the esophagus, but these are not cause for concern. Additionally, there is a small area of scar tissue on the left side of the chest that is likely from a previous injury. | <u>of cancer that has spread from another part of the body (metastatic disease).</u> There is also fluid buildup in the space between the lungs and the chest wall (pleural effusions) on both sides of the body (interstitial lung disease). The radiologist also found some small nodules in the lungs that have not changed since the last exam on [DATE]. | sclerosis of the anterolateral left sixth rib.<br>LLaMA <u>hallucinated</u> possible followup with future scans. |

### User centric evaluation

Table 4 summarizes the expert and laymen evaluation results on 23 radiology reports where we grouped the reports based on the findings and 3 expert radiologists and 3 laymen (people with non-clinical background) evaluated the model generated layman and expert summaries. Given the known variability among the radiologists, we also present the majority opinion for missing information and hallucination. According to the majority opinion, the model didn’t generate any hallucinated information for normal reports and had missing information only for 1 expert summary, while for the metastatic and lung nodule cases model had single hallucinated information and 1 missing information for the layman summary. However linguistic accuracy scored highly for the generated layman summaries 4.18 average Likert scale (high quality). Most hallucinations appeared in “Other Diseases” category which includes findings with relatively smaller representation in the dataset such as fractures. Model sometimes hallucinated the association between rib fracture and pleural effusion while correctly identifying the presence of both the fracture and effusion. On the other hand, the model generated layman summaries obtained an average 63% improvement over the expert summary in terms of understanding scores as assigned by our laymen annotators.

**Table 4:**
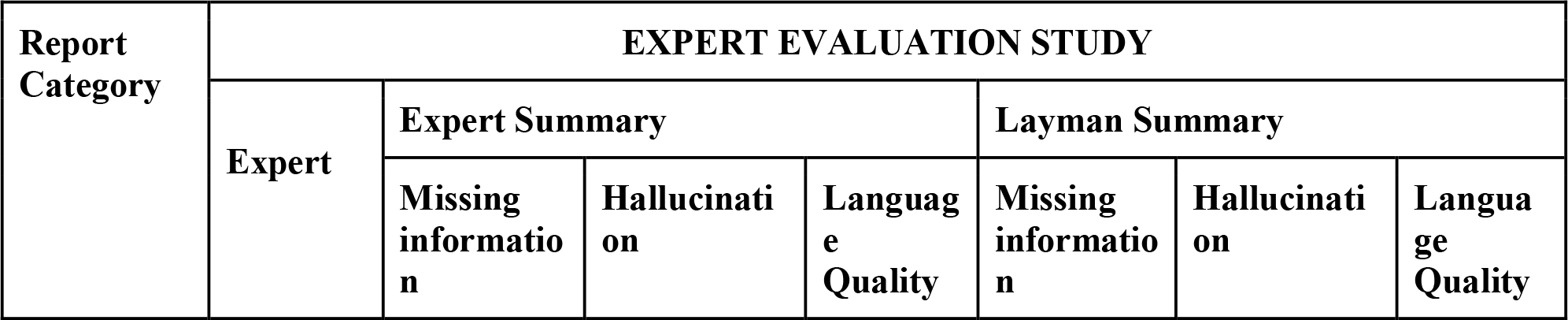
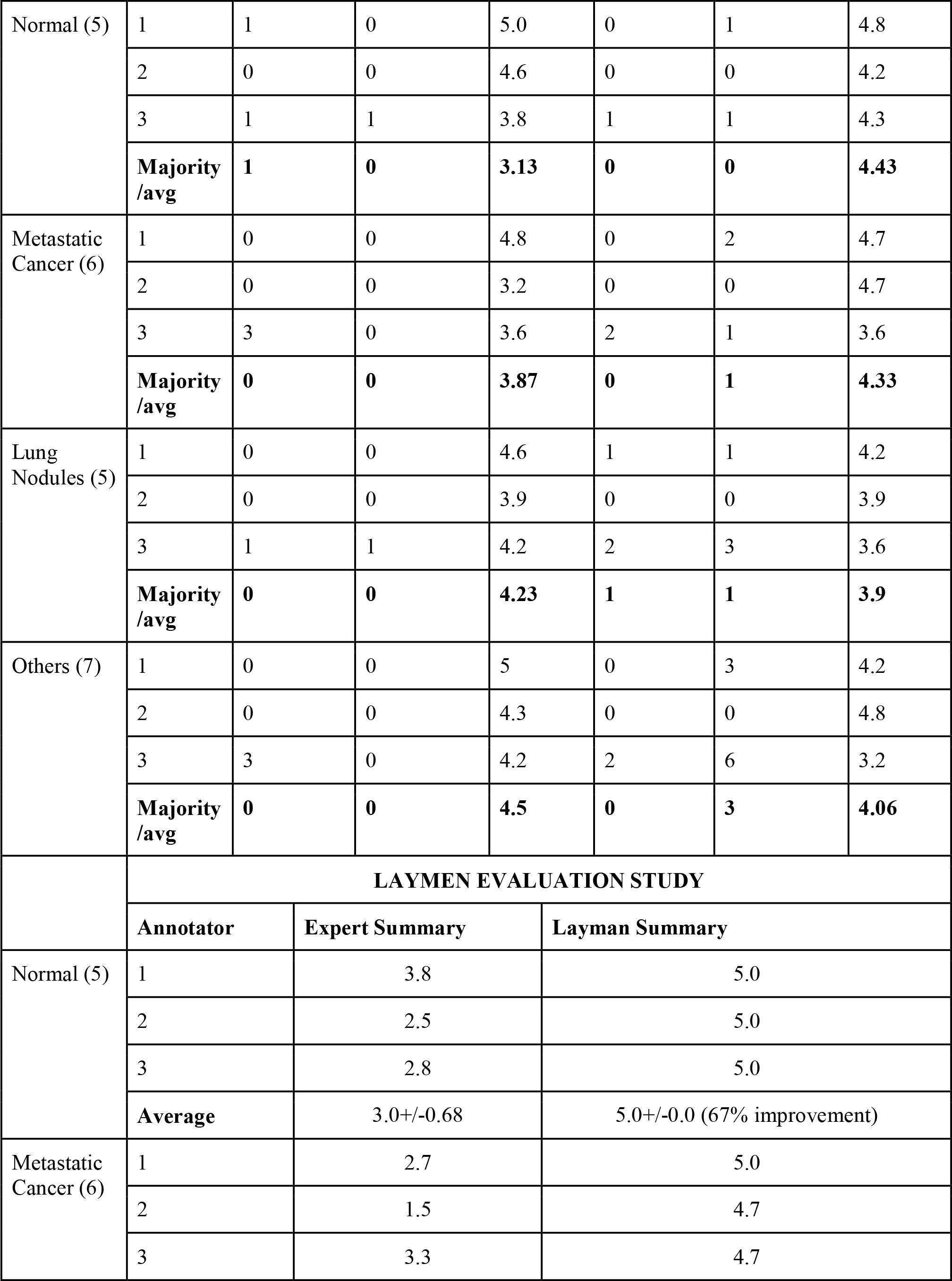
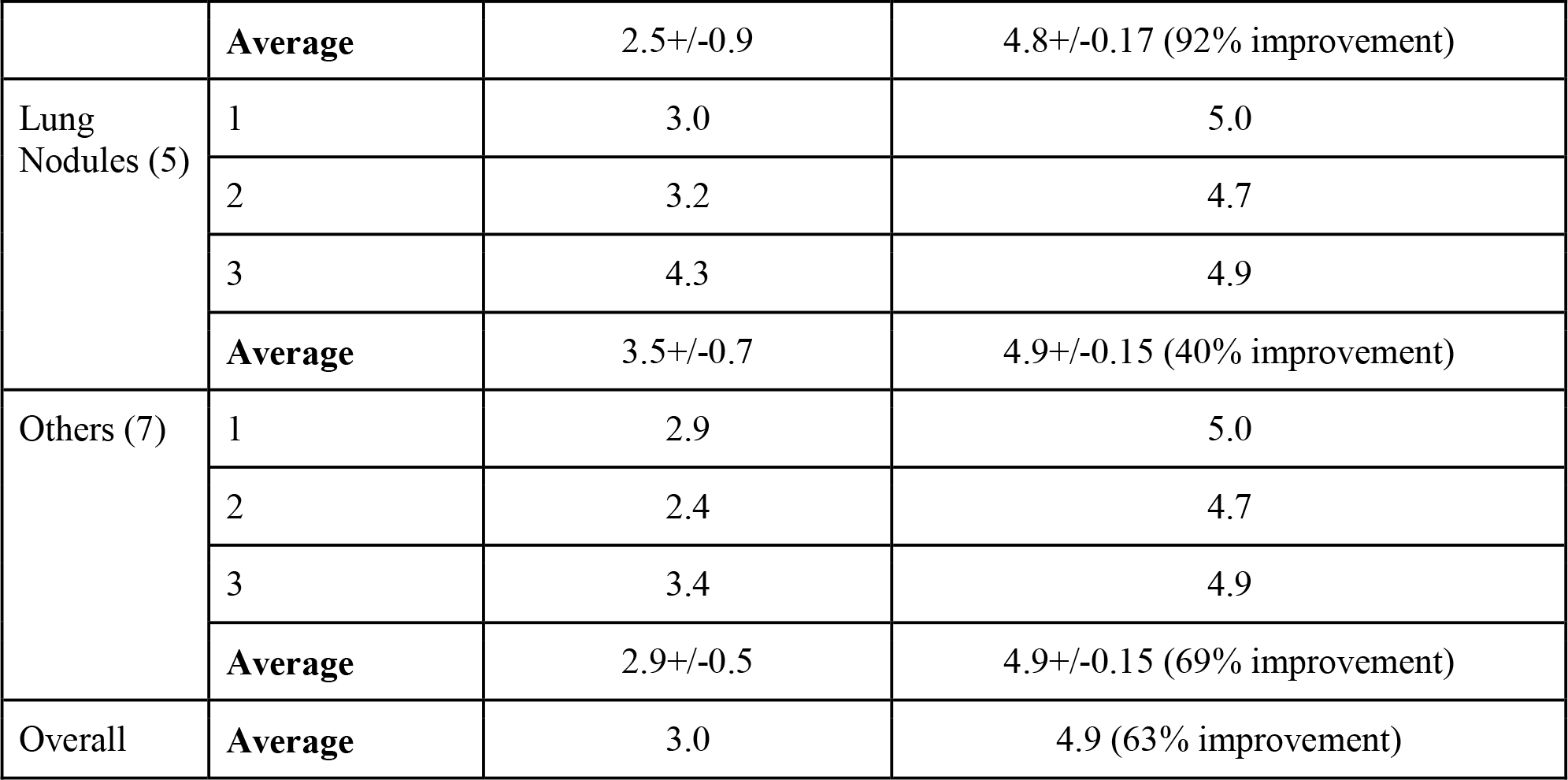
Expert evaluation; Missing information and hallucination columns show number of reports marked with the corresponding issues; Language quality column show average language quality score assigned by each annotator. Non-clinical user evaluation: each column reports average understanding score. Number of reports of each category can be found within parentheses with reports category label (left-most column)

| Report Category | EXPERT EVALUATION STUDY |  |  |  |  |  |  |
| --- | --- | --- | --- | --- | --- | --- | --- |
|  | Expert | Expert Summary |  |  | Layman Summary |  |  |
|  |  | Missing information | Hallucinatio<br>on | Languag<br>e<br>Quality | Missing information | Hallucinati<br>on | Langua<br>ge<br>Quality |
| Normal (5) | 1 | 1 | 0 | 5.0 | 0 | 1 | 4.8 |
|  | 2 | 0 | 0 | 4.6 | 0 | 0 | 4.2 |
|  | 3 | 1 | 1 | 3.8 | 1 | 1 | 4.3 |
|  | **Majority /avg** | **1** | **0** | **3.13** | **0** | **0** | **4.43** |
| Metastatic Cancer (6) | 1 | 0 | 0 | 4.8 | 0 | 2 | 4.7 |
|  | 2 | 0 | 0 | 3.2 | 0 | 0 | 4.7 |
|  | 3 | 3 | 0 | 3.6 | 2 | 1 | 3.6 |
|  | **Majority /avg** | **0** | **0** | **3.87** | **0** | **1** | **4.33** |
| Lung Nodules (5) | 1 | 0 | 0 | 4.6 | 1 | 1 | 4.2 |
|  | 2 | 0 | 0 | 3.9 | 0 | 0 | 3.9 |
|  | 3 | 1 | 1 | 4.2 | 2 | 3 | 3.6 |
|  | **Majority /avg** | **0** | **0** | **4.23** | **1** | **1** | **3.9** |
| Others (7) | 1 | 0 | 0 | 5 | 0 | 3 | 4.2 |
|  | 2 | 0 | 0 | 4.3 | 0 | 0 | 4.8 |
|  | 3 | 3 | 0 | 4.2 | 2 | 6 | 3.2 |
|  | **Majority /avg** | **0** | **0** | **4.5** | **0** | **3** | **4.06** |
|  | **LAYMEN EVALUATION STUDY** |  |  |  |  |  |  |
|  | **Annotator** | **Expert Summary** |  | **Layman Summary** |  |  |  |
| Normal (5) | 1 | 3.8 |  | 5.0 |  |  |  |
|  | 2 | 2.5 |  | 5.0 |  |  |  |
|  | 3 | 2.8 |  | 5.0 |  |  |  |
|  | **Average** | 3.0+/-0.68 |  | 5.0+/-0.0 (67% improvement) |  |  |  |
| Metastatic Cancer (6) | 1 | 2.7 |  | 5.0 |  |  |  |
|  | 2 | 1.5 |  | 4.7 |  |  |  |
|  | 3 | 3.3 |  | 4.7 |  |  |  |

|  |  |  |  |
| --- | --- | --- | --- |
|  | **Average** | 2.5+/-0.9 | 4.8+/-0.17 (92% improvement) |
| Lung Nodules (5) | 1 | 3.0 | 5.0 |
|  | 2 | 3.2 | 4.7 |
|  | 3 | 4.3 | 4.9 |
|  | **Average** | 3.5+/-0.7 | 4.9+/-0.15 (40% improvement) |
| Others (7) | 1 | 2.9 | 5.0 |
|  | 2 | 2.4 | 4.7 |
|  | 3 | 3.4 | 4.9 |
|  | **Average** | 2.9+/-0.5 | 4.9+/-0.15 (69% improvement) |
| Overall | **Average** | 3.0 | 4.9 (63% improvement) |

## DISCUSSION

In this work, we presented a domain-specific fine-tuning of a LLM-based model with noisy groundtruth to generate a patient-centric summary of radiology report findings with the goal of enhancing patients’ understanding of these reports. Patients’ inability to understand their personal medical and clinical status is a critical issue limiting adherence to treatment plans and follow-up appointments. However, no groundtruth dataset - a set of reports with multiple versions of summaries for patients with different levels of knowledge of medical jargon, exist. Given the recent trend of the use of extremely large language models (10’s to 100’s of billion parameters) as zero-shot learners, we used LLaMA to generate layman summaries while using actual impression sections as expert level summaries for training. However, ~15% of layman summaries generated by LLaMA needed to be filtered out because of obvious mistakes, indicating that simple use of LLMs for this task is not sufficient. We fine-tuned a relatively “smaller” LLM - T5 (770m) for the specific task of patient sensitive summary generation which can change language of the summary based on patients’ clinical knowledge - layman or expert. Even though the summarizer model is based on the T5 model with less trainable (~770 million) parameters, the pretrained model suffered severe overfitting for the finetuning unless manual effort was applied to curate ~4K clean training samples from the LLaMA generated noisy groundtruth. Larger models may require even more manual effort to curate larger training sets. This experiment provided motivation to use appropriately sized models for the given task, especially when task-specific finetuning was being performed.

The user evaluation study done by expert radiologists indicates a lack of consensus among experts on what should be reported to the patient. Annotating radiologists disagree on the quality of several model-generated reports. Interestingly, radiologists sometimes even disagree with the reporting made in the original report. For example, the original impression contains “*Prominence of the ascending aorta at 3*.*5 cm*” indicating enlarged aorta which was included in the summary generated by the model. One of the annotating radiologists pointed out that the threshold for aorta enlargement is 4cm, hence, the aorta was not technically enlarged but was reported in an ambiguous manner. In some cases, the model seemed to be adding information about common causes for some findings. For example, the model generated summary included the phrase “*small amount of fluid around the heart, which is likely caused by an infection or inflammation*”. One of the annotating radiologists considered this form of reporting as “hallucination” because while likely correct, the reason was never explicitly mentioned in the original report. An example of disagreement in annotating radiologists’ opinion regarding what should be included in a summary generated for a patient is the exact size of the nodule. While two annotators considered summaries reporting the growth trend of the nodule (growing or stable) sufficient, one annotator considered such cases as having “missing information”. When the report only mentioned unchanged nodes, the model-generated layman summary only stated that no new issues or problems were detected. Some annotators considered this as an incomplete description while others agreed with the model.

Given these disagreements between annotators, we relied on majority opinion for final assessment of model-generated summaries. Among 23 expert summaries, only one case was agreed upon as having missing information where lymphangitic spread of carcinoma was considered a possibility by the reporting radiologist because of the history of cancer. However, the study is designed in such a way that only the finding section was supplied from the original reports for consistency and the model was not provided with the history section of the report, and consequently, missed this possibility. One layman summary was annotated as having missing information based on majority opinion. In that summary, the model failed to describe all indeterminate nodules in the summary. Relatively larger number (5) of layman summaries were considered to have hallucinated information. Model seems to do relatively poorly while generating layman summaries for fractures (categorized under Other Disease). This may be the result of relatively small representation (~10%) of such reports in the training set. While stable exams and metastatic disease also have similar representation in the dataset, their summarization is quite straightforward. In both cases, the patient needs to know their disease is unchanged, or their cancer has now spread. In case of fractures, there are many important pieces of information including anatomical location, severity, and any resulting complications (e.g., joint effusion/hemarthrosis). In another case, the model hallucinated “new spots on liver” when the report mentioned hepatic metastasis. Note that the layman summary generation was trained on noisy groundtruth generated by LLaMA model. Even after manual filtering, it is possible that the training data contains small amounts of discrepancies resulting in limitations on model’s performance for layman summary generation. Even under these circumstances, our non-expert (layman with no medical training) reported on average 63% improvement in their understanding reports when presented with model-generated layman summaries compared to expert summaries, thus proving potential for improvement in patients’ understanding of their clinical status through the use the proposed model.

Comparison between the performance of the proposed model and the zero-shot performance of LLaMA for layman summarization establishes the superiority of our much smaller model both in terms of correctness and lack of hallucination. LLaMA often hallucinated that the report was for chest X-ray instead of chest CT exams and sometimes even hallucinated several abdominal findings just at the mention of a concurrent abdominal MRI in the findings section of the chest CT report. In some cases, LLaMA ignored lung nodules. Our model also sometimes hallucinated the reason for abnormal mass or nodule, even though the model clearly conveyed the uncertainty of such statements.

## CONCLUSION

We designed a first-of-its-kind radiology report summarization model conditioned upon patient clinical knowledge. Our evaluation experiments clearly indicate the limitation of using off-the-shelf LLM as zero-shot learners for this task. On the other hand, noisy ground truth curated from off-the-shelf LLM was successfully used to train a relatively smaller summarizer model which outperformed much larger LLMs for the given task. Two-pronged user study evaluated model generated summaries from points of view of both laymen and experts. Model generated layman summaries were 63% more understandable to laymen. Majority opinion of experts found only a handful of cases with missing or hallucinated information in model generated summaries. In general, experts evaluated expert-level summaries to be better than layman summaries.

## Data Availability

All data produced in the present study are available upon reasonable request to the authors

## Notes

### Competing Interest Statement

The authors have declared no competing interest.

### Funding Statement

This study did not receive any funding

### Author Declarations

Ethical approval granted by Internal Review Board (IRB) of Mayo Clinic.

## REFERENCES

[1] T. Martin-Carreras, T. S. Cook, and C. E. Kahn Jr, “Readability of radiology reports: implications for patient-centered care,” Clin. Imaging, vol. 54, pp. 116–120, 2019.

[2] J. Domingo et al., “Preventing delayed and missed care by applying artificial intelligence to trigger radiology imaging follow-up,” NEJM Catal. Innov. Care Deliv., vol. 3, no. 4, p. CAT–21, 2022.

[3] T. Mabotuwana, C. S. Hall, J. Tieder, and M. L. Gunn, “Improving quality of follow-up imaging recommendations in radiology,” presented at the AMIA annual symposium proceedings, American Medical Informatics Association, 2017, p. 1196.

[4] T. Mabotuwana et al., “Automated tracking of follow-up imaging recommendations,” Am. J. Roentgenol., vol. 212, no. 6, pp. 1287–1294, 2019.

[5] A. Á.-G. Calvillo, L. C. Kodaverdian, R. Garcia, D. Y. Lichtensztajn, and M. D. Bucknor, “Patient-level factors influencing adherence to follow-up imaging recommendations,” Clin. Imaging, vol. 90, pp. 5–10, 2022.

[6] K. Jeblick et al., “ChatGPT makes medicine easy to swallow: an exploratory case study on simplified radiology reports,” Eur. Radiol., pp. 1–9, 2023.

[7] H. Alkaissi and S. I. McFarlane, “Artificial hallucinations in ChatGPT: implications in scientific writing,” Cureus, vol. 15, no. 2, 2023.

[8] M. Sallam, “ChatGPT utility in healthcare education, research, and practice: systematic review on the promising perspectives and valid concerns,” presented at the Healthcare, MDPI, 2023, p. 887.

[9] A. J. Thirunavukarasu, D. S. J. Ting, K. Elangovan, L. Gutierrez, T. F. Tan, and D. S. W. Ting, “Large language models in medicine,” Nat. Med., vol. 29, no. 8, pp. 1930–1940, 2023.

[10] R. Anhang Price et al., “Examining the role of patient experience surveys in measuring health care quality,” Med. Care Res. Rev., vol. 71, no. 5, pp. 522–554, 2014.

[11] O. Alfarghaly, R. Khaled, A. Elkorany, M. Helal, and A. Fahmy, “Automated radiology report generation using conditioned transformers,” Inform. Med. Unlocked, vol. 24, p. 100557, 2021.

[12] S. Dai, Q. Wang, Y. Lyu, and Y. Zhu, “BDKG at MEDIQA 2021: System report for the radiology report summarization task,” presented at the Proceedings of the 20th Workshop on Biomedical Language Processing, 2021, pp. 103–111.

[13] Y. Zhang, D. Y. Ding, T. Qian, C. D. Manning, and C. P. Langlotz, “Learning to summarize radiology findings,” ArXiv Prepr. ArXiv180904698, 2018.

[14] B. Gundogdu et al., “Customized impression prediction from radiology reports using bert and lstms,” IEEE Trans. Artif. Intell., 2021.

[15] X. Cai, S. Liu, J. Han, L. Yang, Z. Liu, and T. Liu, “Chestxraybert: A pretrained language model for chest radiology report summarization,” IEEE Trans. Multimed., 2021.

[16] Y. Zhang, D. Merck, E. B. Tsai, C. D. Manning, and C. P. Langlotz, “Optimizing the factual correctness of a summary: A study of summarizing radiology reports,” ArXiv Prepr. ArXiv191102541, 2019.

[17] Z. Sun et al., “Evaluating GPT-4 on impressions generation in radiology reports,” Radiology, vol. 307, no. 5, p. e231259, 2023.

[18] C. Ma et al., “ImpressionGPT: an iterative optimizing framework for radiology report summarization with chatGPT,” ArXiv Prepr. ArXiv230408448, 2023.

[19] J. Wei et al., “Finetuned Language Models are Zero-Shot Learners,” presented at the International Conference on Learning Representations, 2021.

[20] T. Kojima, S. S. Gu, M. Reid, Y. Matsuo, and Y. Iwasawa, “Large language models are zero-shot reasoners,” Adv. Neural Inf. Process. Syst., vol. 35, pp. 22199–22213, 2022.

[21] M. F. Naeem et al., “I2MVFormer: Large Language Model Generated Multi-View Document Supervision for Zero-Shot Image Classification,” presented at the Proceedings of the IEEE/CVF Conference on Computer Vision and Pattern Recognition, 2023, pp. 15169– 15179.

[22] J. Guo et al., “From Images to Textual Prompts: Zero-shot Visual Question Answering with Frozen Large Language Models,” presented at the Proceedings of the IEEE/CVF Conference on Computer Vision and Pattern Recognition, 2023, pp. 10867–10877.

[23] W. Huang, P. Abbeel, D. Pathak, and I. Mordatch, “Language models as zero-shot planners: Extracting actionable knowledge for embodied agents,” presented at the International Conference on Machine Learning, PMLR, 2022, pp. 9118–9147.

[24] S. Chen et al., “Use of artificial intelligence chatbots for cancer treatment information,” JAMA Oncol., vol. 9, no. 10, pp. 1459–1462, 2023.

[25] I. Banerjee et al., “Natural Language Processing Model for Identifying Critical Findings—A Multi-Institutional Study,” J. Digit. Imaging, vol. 36, no. 1, pp. 105–113, 2023.

[26] H. Touvron et al., “Llama: Open and efficient foundation language models,” ArXiv Prepr. ArXiv230213971, 2023.

[27] C. Raffel et al., “Exploring the limits of transfer learning with a unified text-to-text transformer,” J. Mach. Learn. Res., vol. 21, no. 1, pp. 5485–5551, 2020.

[28] T. Nemoto and D. Beglar, “Likert-scale questionnaires,” presented at the JALT 2013 conference proceedings, 2014, pp. 1–8.

[29] R. Luo et al., “BioGPT: generative pre-trained transformer for biomedical text generation and mining,” Brief. Bioinform., vol. 23, no. 6, p. bbac409, 2022.

